# Self-identification by Sexual and Gender Minorities as a Crucial Aspect of Inclusion in a Study of Adverse Childhood Experiences in Haiti

**DOI:** 10.1101/2023.09.24.23295796

**Authors:** Joseph Heger, Phycien Paul, Caleb Jean-Baptiste, Elmondo Odans, Abraham A. Salinas-Miranda, Erin Lynch, Emily Stevens, Guitele J. Rahill, Manisha Joshi

**Affiliations:** School of Social Work, College of Behavioral and Community Sciences (CBCS) University of South Florida (USF), Florida, United States; Organisation pour la Rénovation de la zone de Cite Soleil (Organization for the Renovation of the Cité Soleil Zone) (OREZON Cité Soleil), Port-au-Prince, Haiti; University Of South Florida (USF), College of Public Health, Tampa, Florida (FL), United States of America (USA)

**Keywords:** Adverse childhood experiences, LGBTQ, gender minorities, Haiti, equity

## Abstract

Research studies are inevitably affected by contextual norms, local ideologies, social mores, and self-identification in addition to extant policies and laws. Self-identification is often emotionally laden and can be constrained by fear of reprisal if one’s self-identification departs from acceptable and affirmed norms. Such complexities can hinder measurement efforts and results from established measures can fail to yield accurate and thus useful data. Studies of Adverse Childhood Experiences (ACEs) are important but understudied in Haiti. We investigated ACEs in urban Haiti using the ACE International Questionnaire (ACE-IQ). Descriptive statistics revealed: (1) A relationship between participants’ self-identification and sex recorded as observed by interviewers, and (2) Prevalence of ACEs by self-reported identity. Roughly half participants self-identified as ’Woman’ (56%; n = 380), 39% as ’Man’ (n = 265), 2% as ’Homosexual’ (n = 13), and 2% as ’Lesbian’ (n =14). Contradictions emerged between participant self-identification and interviewers’ record of sex: 98% self-identified women were observed/recorded as ’Female’, one was observed/recorded as ’Male’, and there was no record for five. Similarly, 95% self-identified men were observed/recorded as ’Male’, six as ’Female’, two as ’Other’, and there was no record for five. Although Sexual and Gender Minorities (SGM) constituted a small subsample (n = 27), their responses underscored misclassification and disproportionate burdens of physical, emotional, and sexual abuse, family dysfunction, and other forms of interpersonal violence. Conducting ACE-IQ studies in Haiti that provide inclusive categories of self-identification may more equitably capture unique ACEs of all Haitians.

---

Research studies of biopsychosocial functioning are inevitably affected by contextual norms, local ideologies, social mores, and self-identification, in addition to extant policies and laws. Self-identification is often emotionally laden and can be constrained by fear of reprisal if one’s self-identification departs from acceptable and affirmed norms. Such complexities can hinder measurement efforts and results from established measures can fail to yield accurate and thus useful data. Studies of adverse childhood Experiences (ACEs) are important but understudied in Haiti. We investigated ACEs in urban Haiti using the ACE International Questionnaire (ACE-IQ).

The need for inclusive and equitable research as a moral and ethical obligation has increased with globalization (Patel & Farmer, 2020; Osel et al., 2022). Central to inclusion and equity in research is consideration of how individuals and groups self-identify and inclusion of underrepresented individuals who reside in Lower-Middle Income Countries (LMIC) such as Haiti (Hamadeh et al., 2021).

The present work summarizes lessons learned while conducting a larger study of ACEs, exposure to neighborhood violence, and behavioral outcomes of violence exposure in Haiti. Within that study, a specific aim was to maximize representation of typically excluded Sexual and Gender Minorities (SGM), especially those who reside in Cité Soleil, a zone that is disproportionately impacted by extreme poverty and interpersonal violence. This aim arose from a previous study in Haiti which reported that transwomen, a subpopulation of SGM, are prone to the same Gender-Based Violence (GBV), rapes, and other forms of violence as individuals who were assigned female at birth (Rahill et al., 2019).

## Haiti

Haiti has a long history of trauma stemming from both internal and external violence. Indeed, Haitians’ experiences of interpersonal violence began in slavery and continued when former Haitian slaves gained their independence from French forces (Charles, 20021). Moreover, “disjunctures in the post-revolutionary governance of Haiti created conditions of precarity, vulnerability, and pre-empted possibilities for development” (Charles, 2021, p.465). Indeed, Haiti’s independence was followed by centuries of interpersonal violence, including the recent assassination of its president (Sanon & Cotto, 2022). In recent years, Haiti earned the dubious recognition of being the most kidnapping-impacted nation in the world (O’Connell, 2021; United States Department of State, n.d.). Recently, conversations regarding the need for a multinational force to quell the current violence and restore order and safety to the populace have been in the news (Blaise, 2023; Human Rights Watch [HRW], 2023).

GBV is one form of interpersonal violence that is commonplace in Haiti. Interpersonal violence includes physical, sexual, domestic, community, and psychological violence – and occurs disproportionately against persons assigned female at birth, including transgendered girls and women (HRW, 2021; Rahill et al., 2018). GBV in Haiti also comprises exclusion of SGM in research and intervention.

### Haiti’s Cité Soleil

Cité Soleil, phonetically *See’tay So’Lay*, is situated in Haiti’s capital of Port-au-Prince and has roughly 400,000 residents crowded in less than nine square miles (Buschschlüter, 2014). Cité Soleil comprises of 30 sectors that are each governed by feared gang leaders (Buschschlüter, 2022; Rahill et al., 2019). Nevertheless, its non-criminal residents are peace-loving and intelligent who treasure education, and long for safety and security (Torgan, 2019).

In Cité Soleil, a complex interaction of extreme poverty, structural and community violence, experience of traumatic events across the lifespan, sleep disruptions, infectious diseases, and youths’ involvement in gang warfare result in a life expectancy of 45-50 years, lower than Haiti’s 64 years (Buschschlüter, 2014; World Population, 2022). Thus, the zone’s population is quite young.

For Cité Soleil SGM, experiences of interpersonal and community violence are increased due to religious beliefs that disparage non-heteronormative gender expression, local norms that minimize women’s worth, widespread substance abuse, and easy access to weapons (Durban-Albrecht, 2017; Joshi et al., 2021). In addition, a 2020 penal code in Haiti prescribes imprisonment for crimes against SGM, falls short of removing steep financial penalties for same-sex marriage, and prohibits a certificate of good standing to non-cisgender and non-heterosexual individuals (HRW, 2021). It follows that the health and well-being of SGM are important topics for research and intervention. Researchers have documented intentional and unintentional exclusion of SGM in studies that are pertinent to their health and wellbeing (Rahill et al., 2018). Lacking has been knowledge on ACEs and behavioral outcomes of violence exposure in SGM populations in Haiti. However, for Haitian SGM, whose identities and social positions in the local, regional, and global environments constitute explicit risks to their safety and health, knowledge of ACEs is crucial.

## Adverse Childhood Experiences (ACEs)

ACEs include experiences of neglect, emotional, physical, and sexual abuse, exposure to substance abuse, and witnessing domestic, peer, community, and gang violence (Centers for Disease Control and Prevention [CDC], 2021). ACEs give rise to poor biopsychosocial outcomes across the life cycle. Indeed, the World Health Organization (WHO) in 2020 noted,

> …considerable and prolonged stress in childhood has life-long consequences…can disrupt early brain development and compromise functioning of the nervous and immune systems…can lead to serious problems such as alcoholism, depression, eating disorders, unsafe sex, HIV/AIDS, heart disease, cancer…

ACEs have been studied in Low-Income Countries (LIC)/LMIC such as Nigeria (Kazeem, 2015), Malawi (Kidman et al., 2020), and similar settings, using the ACE-IQ, detailed below. Lacking are studies that use the ACE-IQ to investigate ACES and SGM in Haiti.

## Sexual and Gender Minorities (SGM)

According to the National Institutes of Health (2019), SGM are individuals:

> … who identify as lesbian, gay, bisexual, asexual, transgender, Two-Spirit, queer, and/or intersex. Individuals with same-sex or -gender attractions or behaviors and those with a difference in sex development …those who do not self-identify with one of these terms but whose sexual orientation, gender identity or expression, or reproductive development is characterized by non-binary constructs of sexual orientation, gender, and/or sex.

### Key Concepts

#### Sex and Sexuality

Sex is assigned at birth based on chromosomes and genitals: Individuals with XX chromosomes, ovaries, and estrogen are labelled female while individuals with XY chromosomes, testes, and testosterone are labelled male (National Academies of Sciences, Engineering, and Medicine [NASEM], 2022). This classification is based on reproductive anatomy, where historically the male/female or man/woman binary has been the norm (NASEM, 2022). Nevertheless, 1 in 2,000 people are born with anatomy and/or chromosomes that do not fall into either of the latter categories, and who arbitrarily are medically classified as intersex (NASEM, 2022). Sexuality is “one’s romantic, sexual, and emotional attraction to others”; a person may be sexually but not romantically drawn to a certain gender (Jourian, 2015, p. 14).

There are approximately two dozen intersex traits, which justifies subjective self-identification. One intersex class comprises individuals with Complete Androgen Insensitivity Syndrome (CAIS) (Singh & Ilyayeva, 2022). People with CAIS, who often are female presenting, have a vagina and undescended testes, and have XY chromosomes, are often pressured to classify as either a man or woman, and frequently struggle with gender identity (Singh et al., 2022). Some individuals with CAIS are even subjected to purported *corrective* procedures to place them medically closer to one side of the binary. Yet, the need of self-identification by SGM is crucial in recognition of SGM dignity and autonomy and is a matter of social justice and equity.

#### Gender

Gender is central to understanding health inequities and their origins (WHO, 2020a). Although often conflated, the concepts of sex, gender, gender identity, gender expression, transgender, and sexuality must be disentangled for equitable research and treatment of SGM (NASEM, 2022). Gender is socially constructed and is historically defined based on norms, roles, and behaviors that a given society deems appropriate, based on an individual’s birth sex (WHO, 2021). Gender *roles* typically refer to what society expects each gender to do at home, work, and other settings (WHO, 2021). Gender *identity* refers to individuals’ experiences and feelings about how they perceive their gender and may or may not coincide with the sex and/or gender assigned at birth (WHO 2021). Gender identities inform how individuals experience their bodies, their manner of dress or decisions to modify the physical structure of their bodies, their sexual orientation, and/or their linguistic and behavioral expressions (National Institutes of Health, 2019). Gender *expression* is the performance and enactment of gender. It is not always consonant with gender presentation or with sexual orientation. For example, an individual who identifies as gender-fluid can be female-presenting and have partners that are homosexual, heterosexual, or pansexual. Importantly, one’s gender expression does not constitute a valid method for categorization for purposes of service provision since a person’s gender expression may not match their gender identity, and sexuality and gender both exist on a continuum (Restar et al., 2021).

Transgender describes individuals whose gender identity or expression diverges from culturally constructed categories for sex and gender. Within the transgender category, there are trans-men, trans-women, and sometimes individuals who identify as nonbinary, who may assume gender roles based on their self-identification (Rahill et al., 2018). Gender norms and roles establish gender relations, and these relations often create hierarchies that result in disadvantages for one or more genders (WHO, 2021).

#### Health Disparities and Haitian SGM

A growing body of knowledge highlights ACEs of SGM and adverse outcomes (Felitti et al., 1998; Hollinsaid et al., 2020; Mustanski et al., 2021; Qureshi et al., 2022; Schnarrs et al., 2022). That literature indicates that SGM bear disproportionate burdens of societal ills and health disparities, including psychological distress, mood disturbances, trauma, cardiovascular disease, anxiety, and others (Andersen & Blosnich, 2013; Lund & Burgess, 2021).

In Cité Soleil, SGM health disparities are linked with sociopolitical conditions, its poor record of human rights, alienation, stigmatization, and experiences of violence. Specifically, Cité Soleil SGM reportedly face bullying, stabbings, beatings, being shot, emotional abuse, and sexual coercion (Rahill et al., 2020; Joshi et al., 2021). Previous studies report that Haitian SGM lack awareness of terminology that would clarify distinctions among sex, gender, gender identity, gender roles and gender expression and, therefore, self-identify using pejorative terms imposed by their heteronormative peers (Rahill et al., 2020). Additionally, HIV prevalence for SGM has increased while Haiti’s prevalence has declined from over 3% to less than 2% (Dévieux et al., 2022).

## Materials and Methods

This cross-sectional, community survey was conducted between November 2020 and January 2021 by Haiti-based colleagues at Organization for the Renovation of the Cité Soleil Zone (OREZON). OREZON is a community social service organization that enjoys the trust and respect of the community. Our research team has collaborated with OREZON in previous research that is pertinent to Cité Soleil residents and that builds organization capacity. OREZON colleagues were certified in the protection of human subjects through Family Health International.

This study follows the WHO’s recommendations for administering the ACE-IQ. We established rapport with key stakeholder organizations, ensured availability of service referrals to participants, and, to maximize linguistic accuracy, we translated and back-translated the ACE-IQ from English to Haitian *Kreyòl*, etc. (WHO, 2018).

OREZON colleagues used venue-based and convenience sampling to recruit 673 volunteer participants across Cité Soleil’s 30 sectors. Upon completion of the informed consent process, adult participants completed the ACE-IQ, sociodemographic questions, and questions based on previous studies in Cité Soleil. Given our objective of maximizing SGM representation, OREXON colleagues intentionally recruited in SGM enclaves of Cité Soleil. One recruited person did not respond to the self-identification question of ‘How do you identify’ and was dropped. Our final sample was 672. At OREZON colleagues’ request, our university signed a data use agreement, following which we obtained the approval of our university’s Institutional Review Board to analyze and interpret the data with them.

### Measures

The WHO in 2022 describes the ACE-IQ as a structured instrument for individuals aged 18 or older, which assesses:

> …some of the most intensive and frequently occurring sources of stress that children may suffer early in life…multiple types of abuse; neglect; violence between parents or caregivers; other kinds of serious household dysfunction such as alcohol and substance abuse; and peer, community, and collective violence…

The ACE-IQ enables: (1) comparison of ACEs across nations, associations between ACEs and health risk behaviors and health outcomes in adult life, advocacy for increased investments to reduce ACEs, development of appropriate prevention programs (WHO, 2020b). Critics of the ACE-IQ address its lack of focus on unique LIC/LMIC concerns but grant that it is credible, valid, and reliable (Pereira, 2019; Rutter, 2021).

The ACE-IQ measures 13 categories of ACEs divided into four dimensions: (1) Abuse (physical, emotional, contact sexual); (2) Parental neglect; (3) Family dysfunction, and (4) Violence outside the household (WHO, 2022). We used the full ACE-IQ, with one adaptation to the first item that requires the interviewer to ‘Record Male/Female as observed.’ We added ‘Other’, to prevent forcing the interviewer to record a participant as either Male or Female when unsure. We also added a question which assessed how participants self-identified in terms of gender and sexual orientation. The response categories included: as a *woman*, *man*, *homosexual*, *lesbian*, and *I have a different identification*. Although we are aware that *woman* and *man* denote sex and *homosexual* and *lesbian* denote sexual orientation, we used these terms based on extant knowledge that contextual nomenclature for non-heteronormative persons in Haiti are typically adopted and used by Cité Soleil SGM (Joshi et al., 2021). Finally, we added 20 sociodemographic questions (age, relationship status, education, employment status, income).

### Analysis

The ACE-IQ has two options for coding, binary (yes, no) or frequency (e.g., many times, few times, one, never). We used the frequency option, to assess differences in ACE severity (WHO, 2022). Descriptive statistics permitted exploration of participants’ socio-demographics, self-identification (gender, sexual orientation), responses to sex as observed and recorded by the interviewer, and differences in ACEs by self-identification categories (i.e., man, woman, homosexual, lesbian, other). Of particular interest was exploration of the overlap (or lack thereof) between interviewers’ record of ‘Sex as observed’ and participants’ self-identification in terms of gender and sexual orientation. Tests of statistical inference were omitted given the small sample sizes for the participants who identified as homosexual (n=13) and lesbian (n=14) (Contextually normed and accepted cover term for non-binary and non-heteronormative persons in Haiti) (Rahill et al., 2019). STATA version 17 was used for analyses.

## Results

### Participant Characteristics

Most of our sample (N=672) was younger than 31 years (n = 463; 72.5%), consistent with previous findings of Cité Soleil’s young population (Buschschlüter, 2014). Three-fourths of the participants had completed less than high school (n = 463; 72.5%), over half (n=384; 58.6%) were unemployed, and more than half (n = 388; 58%) were recorded by the interviewer as female and about two-fifth (n = 256; 38%) as male. The remaining 3% (n=19) were marked as other and data were missing for 1.5% participants (n=10). More than half of participants self-identified as ‘woman’ (n = 380; 56%) and about two-fifth (n = 265; 39%) as a ‘man.’ About 2% self-identified as ‘homosexual’ (n = 13) and another 2% (n = 14) self-identified as ‘lesbian.’

### Relationship Between Sex Recorded as Observed and Self-identification

Among participants who self-identified as woman (n=380), a substantial majority (n = 374; 98.42%) were observed/recorded by the interviewer as female, one as male, and for 5 there was no record. Similarly, among those who self-identified as man (n=265), a substantial majority (n = 252; 95.09%) were observed/recorded as male, six as female, 2 as other, and there was no interviewer record for 5. For the 13 participants who self-identified as homosexual, a majority (n=10) were recorded by the interviewer as other, 2 as male, and 1 as female. Half (n = 7; 50%) of those who self-identified as lesbian (n=14) were recorded by the interviewer as other, 6 as female, and 1 as male.

### Domains of ACE-IQ

Table 1 presents a comprehensive picture of ACE domains by self-identified categories. Although sample sizes for self-identified as homosexual or lesbian were small, the ACE patterns that emerged are noteworthy. Some findings are highlighted below.

**Table 1.**
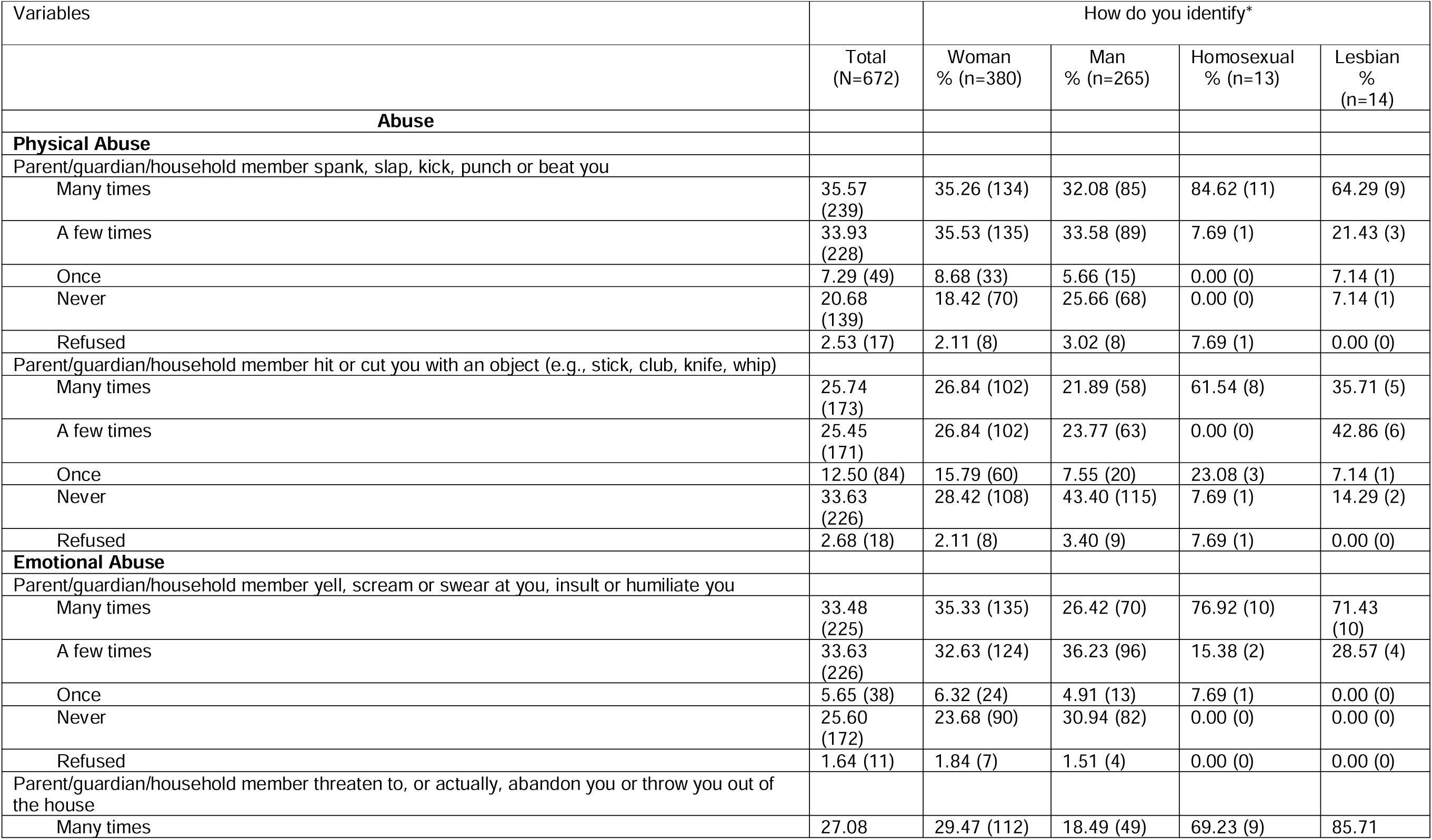
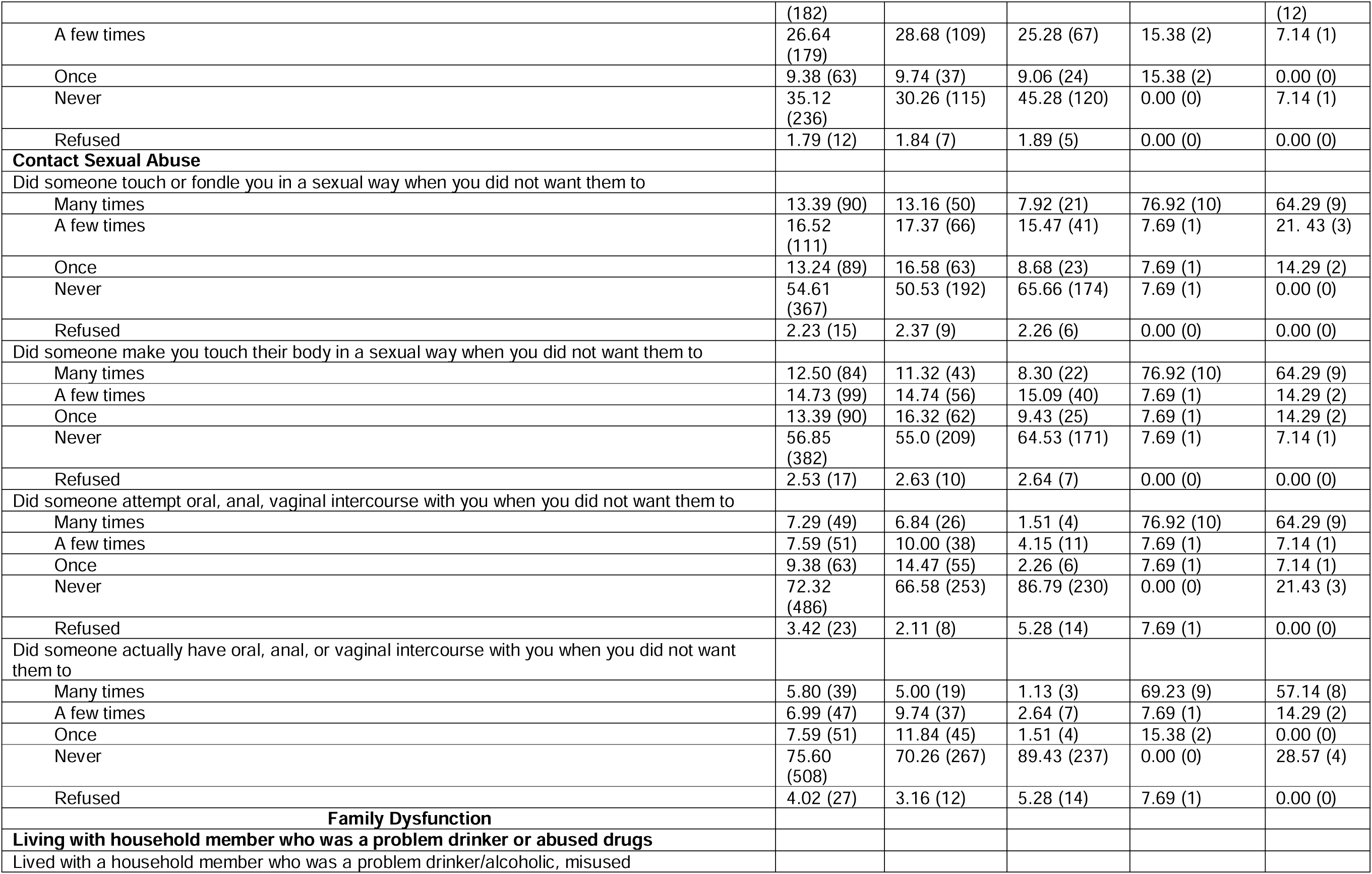
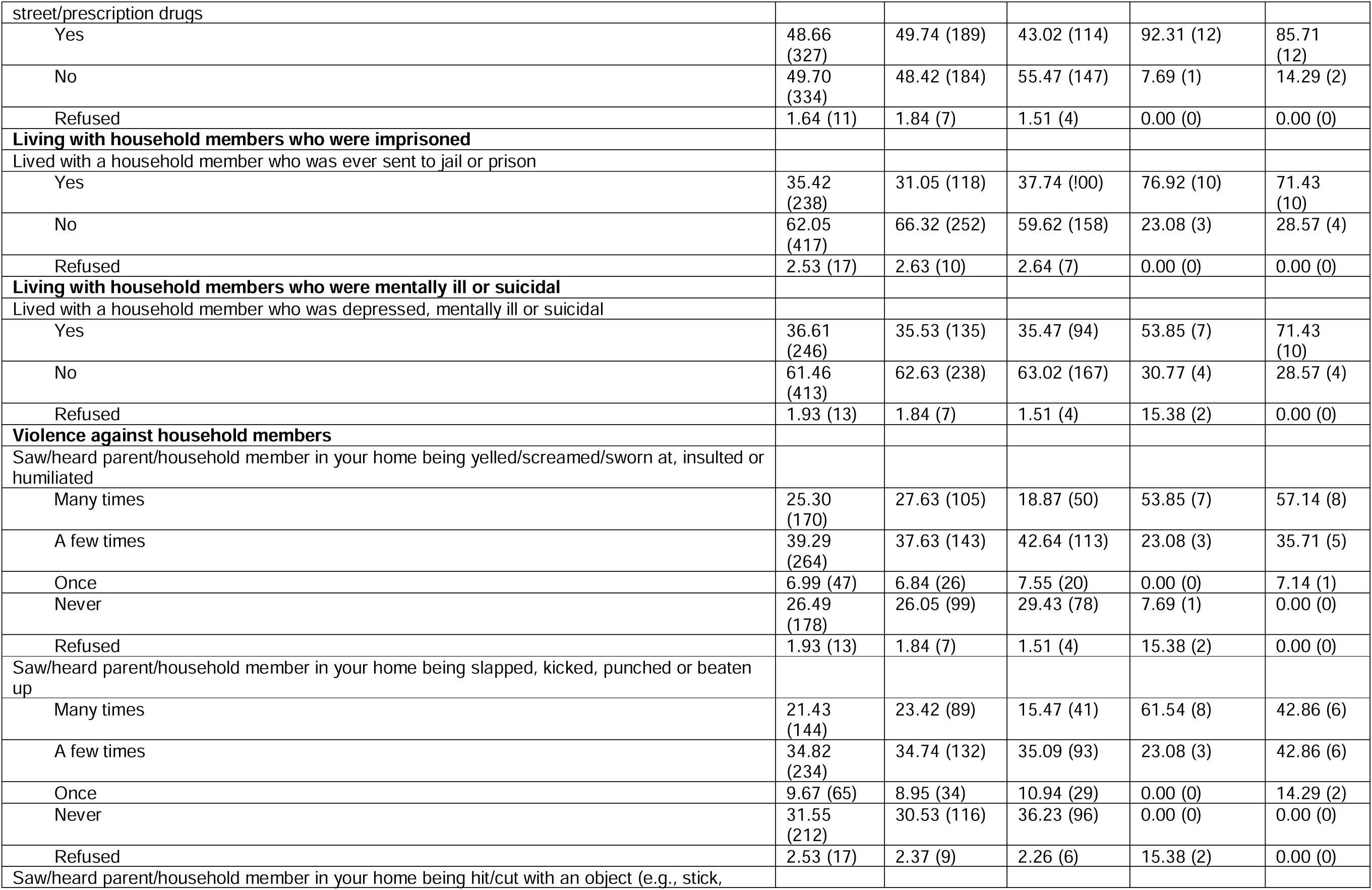
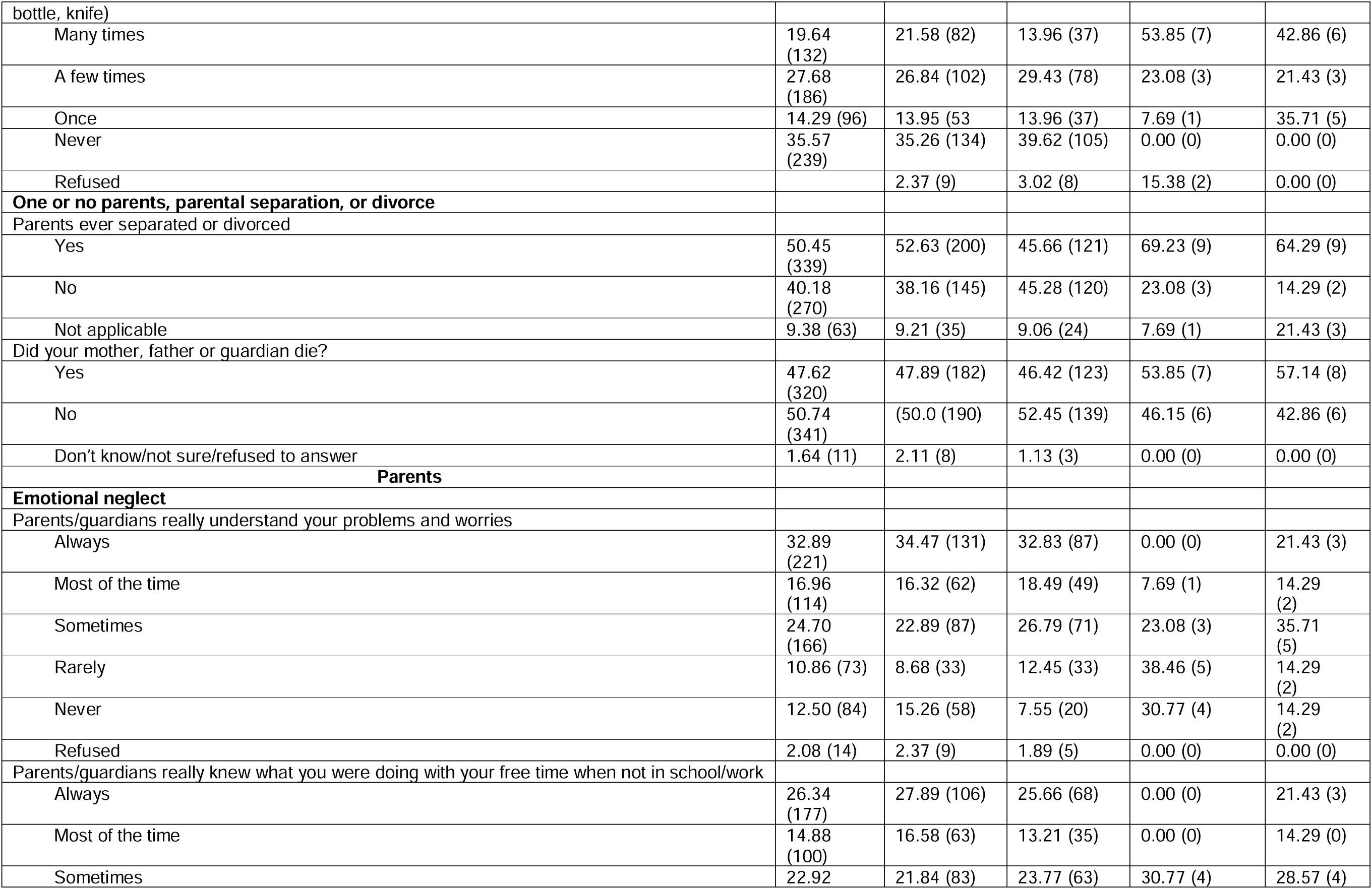
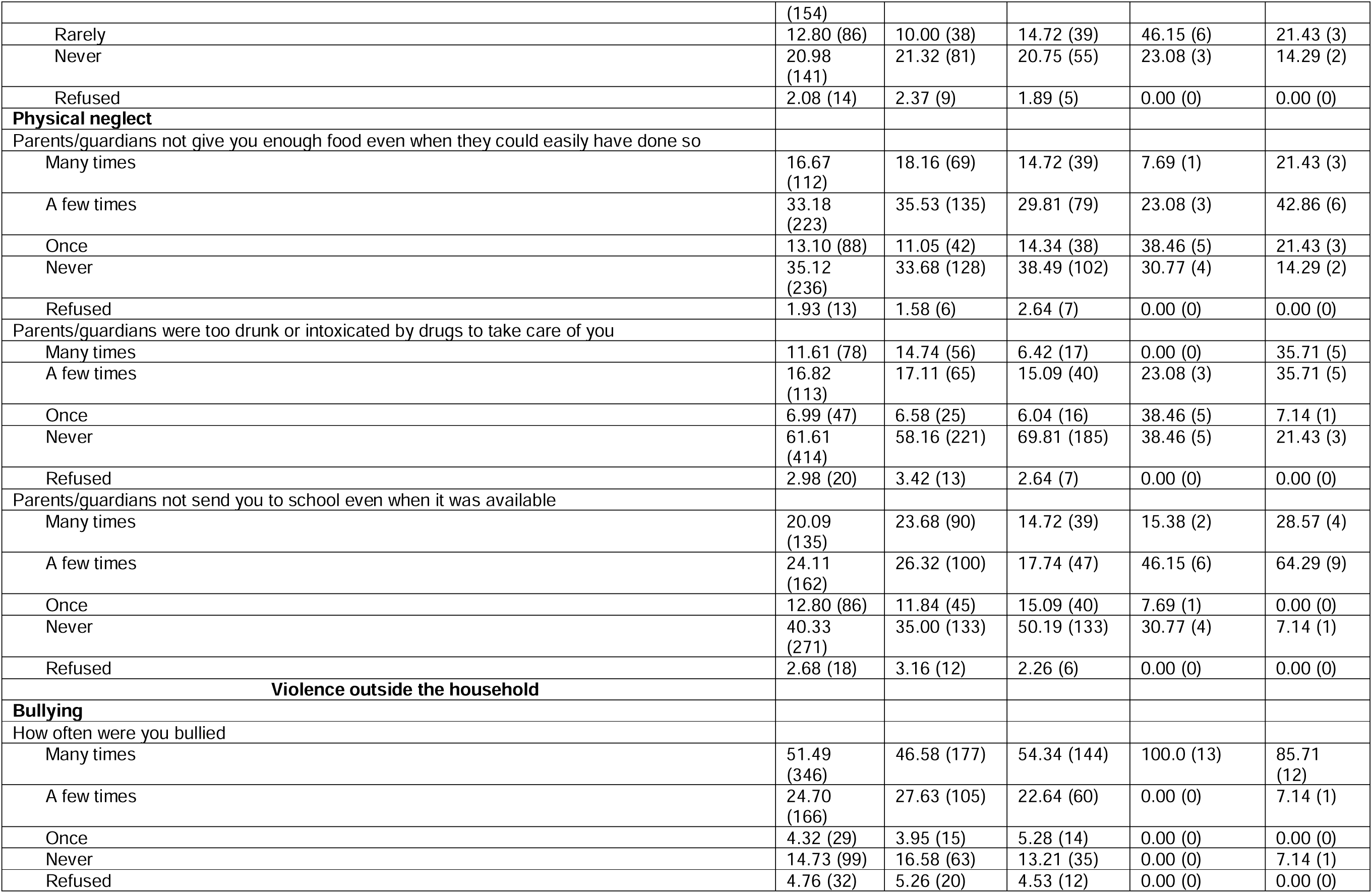
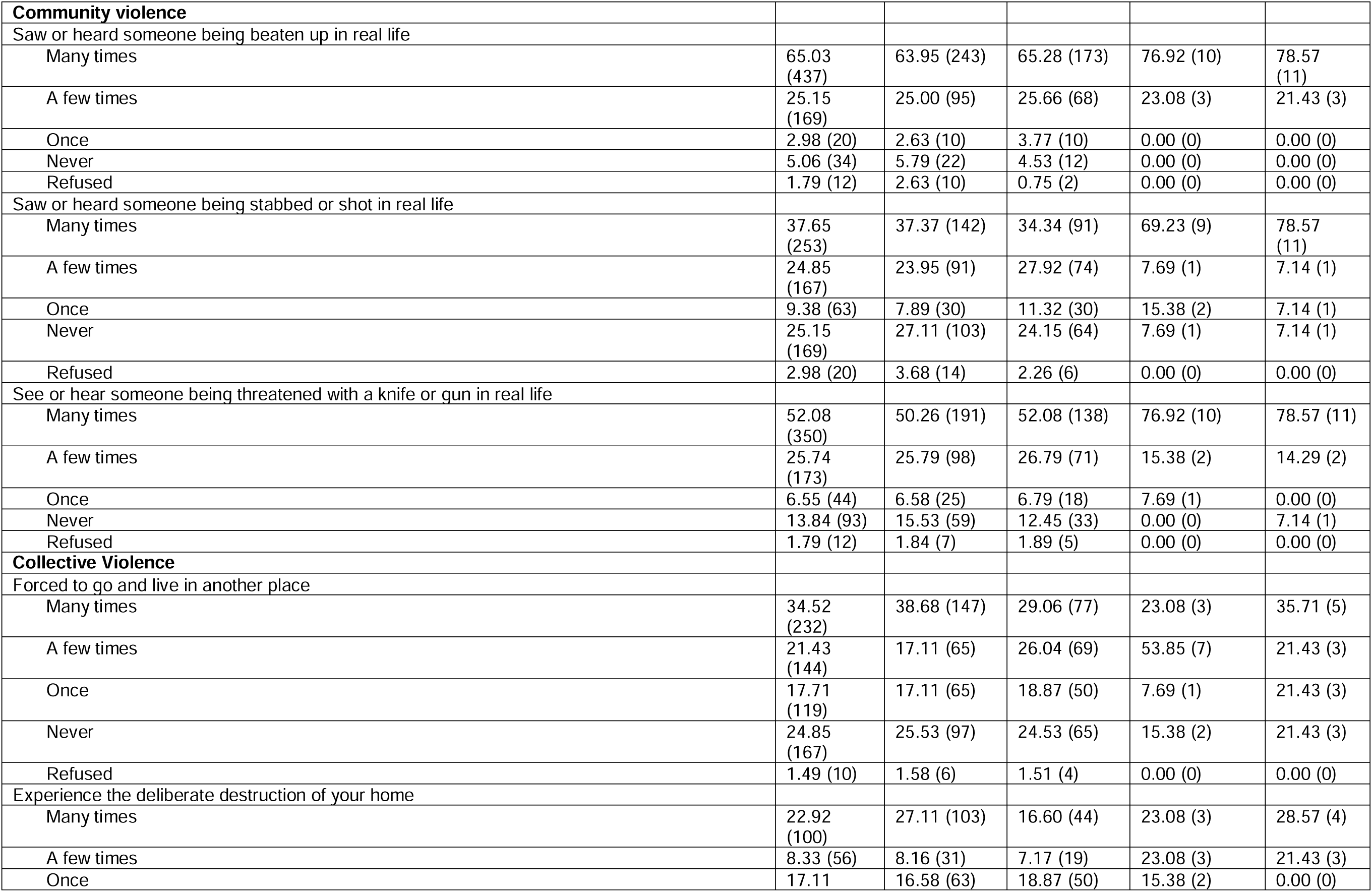
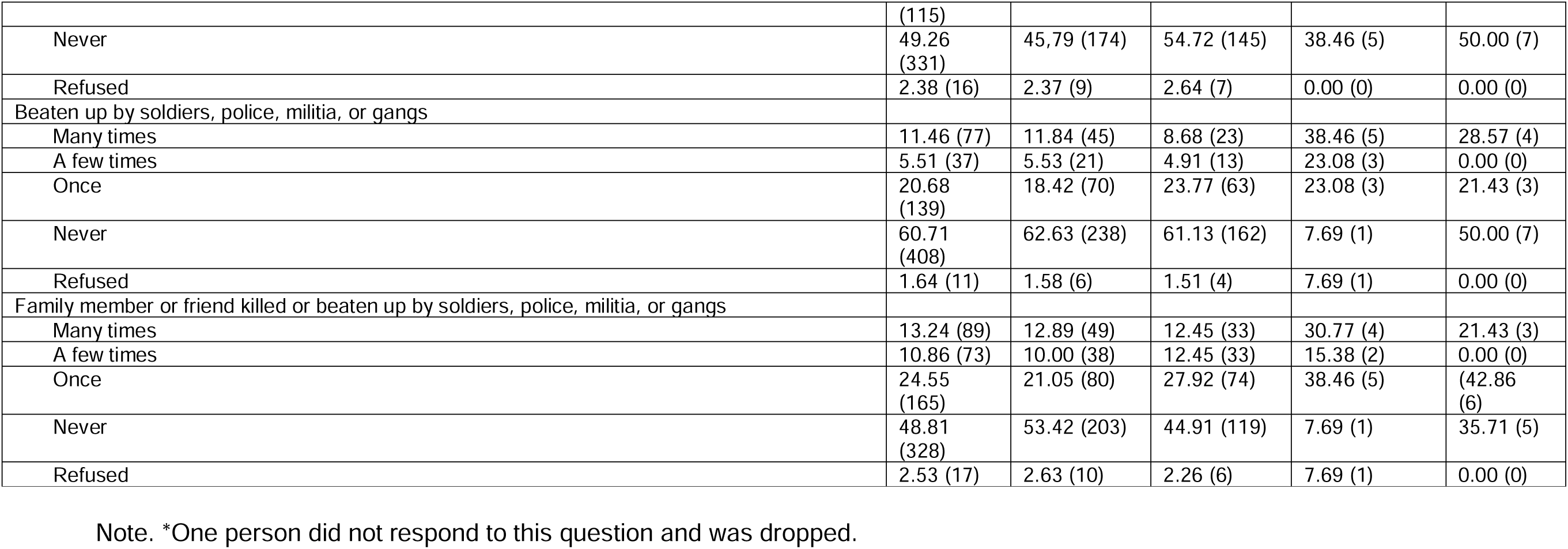
Percentage Prevalence of ACE-IQ Dimensions according to Self-identification in terms of Gender and Sexual Orientation, Cité Soleil Haiti, January-June 2021 (N=672)

| Variables |  | How do you identify* |  |  |  |
| --- | --- | --- | --- | --- | --- |
|  | Total<br>(N=672) | Woman<br>% (n=380) | Man<br>% (n=265) | Homosexual<br>% (n=13) | Lesbian<br>%<br>(n=14) |
| **Abuse** |  |  |  |  |  |
| **Physical Abuse** |  |  |  |  |  |
| Parent/guardian/household member spank, slap, kick, punch or beat you |  |  |  |  |  |
| Many times | 35.57<br>(239) | 35.26 (134) | 32.08 (85) | 84.62 (11) | 64.29 (9) |
| A few times | 33.93<br>(228) | 35.53 (135) | 33.58 (89) | 7.69 (1) | 21.43 (3) |
| Once | 7.29 (49) | 8.68 (33) | 5.66 (15) | 0.00 (0) | 7.14 (1) |
| Never | 20.68<br>(139) | 18.42 (70) | 25.66 (68) | 0.00 (0) | 7.14 (1) |
| Refused | 2.53 (17) | 2.11 (8) | 3.02 (8) | 7.69 (1) | 0.00 (0) |
| Parent/guardian/household member hit or cut you with an object (e.g., stick, club, knife, whip) |  |  |  |  |  |
| Many times | 25.74<br>(173) | 26.84 (102) | 21.89 (58) | 61.54 (8) | 35.71 (5) |
| A few times | 25.45<br>(171) | 26.84 (102) | 23.77 (63) | 0.00 (0) | 42.86 (6) |
| Once | 12.50 (84) | 15.79 (60) | 7.55 (20) | 23.08 (3) | 7.14 (1) |
| Never | 33.63<br>(226) | 28.42 (108) | 43.40 (115) | 7.69 (1) | 14.29 (2) |
| Refused | 2.68 (18) | 2.11 (8) | 3.40 (9) | 7.69 (1) | 0.00 (0) |
| **Emotional Abuse** |  |  |  |  |  |
| Parent/guardian/household member yell, scream or swear at you, insult or humiliate you |  |  |  |  |  |
| Many times | 33.48<br>(225) | 35.33 (135) | 26.42 (70) | 76.92 (10) | 71.43<br>(10) |
| A few times | 33.63<br>(226) | 32.63 (124) | 36.23 (96) | 15.38 (2) | 28.57 (4) |
| Once | 5.65 (38) | 6.32 (24) | 4.91 (13) | 7.69 (1) | 0.00 (0) |
| Never | 25.60<br>(172) | 23.68 (90) | 30.94 (82) | 0.00 (0) | 0.00 (0) |
| Refused | 1.64 (11) | 1.84 (7) | 1.51 (4) | 0.00 (0) | 0.00 (0) |
| Parent/guardian/household member threaten to, or actually, abandon you or throw you out of the house |  |  |  |  |  |
| Many times | 27.08 | 29.47 (112) | 18.49 (49) | 69.23 (9) | 85.71 |
|  | (182) |  |  |  | (12) |
| A few times | 26.64 (179) | 28.68 (109) | 25.28 (67) | 15.38 (2) | 7.14 (1) |
| Once | 9.38 (63) | 9.74 (37) | 9.06 (24) | 15.38 (2) | 0.00 (0) |
| Never | 35.12 (236) | 30.26 (115) | 45.28 (120) | 0.00 (0) | 7.14 (1) |
| Refused | 1.79 (12) | 1.84 (7) | 1.89 (5) | 0.00 (0) | 0.00 (0) |
| **Contact Sexual Abuse** |  |  |  |  |  |
| Did someone touch or fondle you in a sexual way when you did not want them to |  |  |  |  |  |
| Many times | 13.39 (90) | 13.16 (50) | 7.92 (21) | 76.92 (10) | 64.29 (9) |
| A few times | 16.52 (111) | 17.37 (66) | 15.47 (41) | 7.69 (1) | 21.43 (3) |
| Once | 13.24 (89) | 16.58 (63) | 8.68 (23) | 7.69 (1) | 14.29 (2) |
| Never | 54.61 (367) | 50.53 (192) | 65.66 (174) | 7.69 (1) | 0.00 (0) |
| Refused | 2.23 (15) | 2.37 (9) | 2.26 (6) | 0.00 (0) | 0.00 (0) |
| Did someone make you touch their body in a sexual way when you did not want them to |  |  |  |  |  |
| Many times | 12.50 (84) | 11.32 (43) | 8.30 (22) | 76.92 (10) | 64.29 (9) |
| A few times | 14.73 (99) | 14.74 (56) | 15.09 (40) | 7.69 (1) | 14.29 (2) |
| Once | 13.39 (90) | 16.32 (62) | 9.43 (25) | 7.69 (1) | 14.29 (2) |
| Never | 56.85 (382) | 55.0 (209) | 64.53 (171) | 7.69 (1) | 7.14 (1) |
| Refused | 2.53 (17) | 2.63 (10) | 2.64 (7) | 0.00 (0) | 0.00 (0) |
| Did someone attempt oral, anal, vaginal intercourse with you when you did not want them to |  |  |  |  |  |
| Many times | 7.29 (49) | 6.84 (26) | 1.51 (4) | 76.92 (10) | 64.29 (9) |
| A few times | 7.59 (51) | 10.00 (38) | 4.15 (11) | 7.69 (1) | 7.14 (1) |
| Once | 9.38 (63) | 14.47 (55) | 2.26 (6) | 7.69 (1) | 7.14 (1) |
| Never | 72.32 (486) | 66.58 (253) | 86.79 (230) | 0.00 (0) | 21.43 (3) |
| Refused | 3.42 (23) | 2.11 (8) | 5.28 (14) | 7.69 (1) | 0.00 (0) |
| Did someone actually have oral, anal, or vaginal intercourse with you when you did not want them to |  |  |  |  |  |
| Many times | 5.80 (39) | 5.00 (19) | 1.13 (3) | 69.23 (9) | 57.14 (8) |
| A few times | 6.99 (47) | 9.74 (37) | 2.64 (7) | 7.69 (1) | 14.29 (2) |
| Once | 7.59 (51) | 11.84 (45) | 1.51 (4) | 15.38 (2) | 0.00 (0) |
| Never | 75.60 (508) | 70.26 (267) | 89.43 (237) | 0.00 (0) | 28.57 (4) |
| Refused | 4.02 (27) | 3.16 (12) | 5.28 (14) | 7.69 (1) | 0.00 (0) |
| **Family Dysfunction** |  |  |  |  |  |
| **Living with household member who was a problem drinker or abused drugs** |  |  |  |  |  |
| Lived with a household member who was a problem drinker/alcoholic, misused |  |  |  |  |  |
| street/prescription drugs |  |  |  |  |  |
| Yes | 48.66<br>(327) | 49.74 (189) | 43.02 (114) | 92.31 (12) | 85.71<br>(12) |
| No | 49.70<br>(334) | 48.42 (184) | 55.47 (147) | 7.69 (1) | 14.29 (2) |
| Refused | 1.64 (11) | 1.84 (7) | 1.51 (4) | 0.00 (0) | 0.00 (0) |
| **Living with household members who were imprisoned** |  |  |  |  |  |
| Lived with a household member who was ever sent to jail or prison |  |  |  |  |  |
| Yes | 35.42<br>(238) | 31.05 (118) | 37.74 (100) | 76.92 (10) | 71.43<br>(10) |
| No | 62.05<br>(417) | 66.32 (252) | 59.62 (158) | 23.08 (3) | 28.57 (4) |
| Refused | 2.53 (17) | 2.63 (10) | 2.64 (7) | 0.00 (0) | 0.00 (0) |
| **Living with household members who were mentally ill or suicidal** |  |  |  |  |  |
| Lived with a household member who was depressed, mentally ill or suicidal |  |  |  |  |  |
| Yes | 36.61<br>(246) | 35.53 (135) | 35.47 (94) | 53.85 (7) | 71.43<br>(10) |
| No | 61.46<br>(413) | 62.63 (238) | 63.02 (167) | 30.77 (4) | 28.57 (4) |
| Refused | 1.93 (13) | 1.84 (7) | 1.51 (4) | 15.38 (2) | 0.00 (0) |
| **Violence against household members** |  |  |  |  |  |
| Saw/heard parent/household member in your home being yelled/screamed/sworn at, insulted or humiliated |  |  |  |  |  |
| Many times | 25.30<br>(170) | 27.63 (105) | 18.87 (50) | 53.85 (7) | 57.14 (8) |
| A few times | 39.29<br>(264) | 37.63 (143) | 42.64 (113) | 23.08 (3) | 35.71 (5) |
| Once | 6.99 (47) | 6.84 (26) | 7.55 (20) | 0.00 (0) | 7.14 (1) |
| Never | 26.49<br>(178) | 26.05 (99) | 29.43 (78) | 7.69 (1) | 0.00 (0) |
| Refused | 1.93 (13) | 1.84 (7) | 1.51 (4) | 15.38 (2) | 0.00 (0) |
| Saw/heard parent/household member in your home being slapped, kicked, punched or beaten up |  |  |  |  |  |
| Many times | 21.43<br>(144) | 23.42 (89) | 15.47 (41) | 61.54 (8) | 42.86 (6) |
| A few times | 34.82<br>(234) | 34.74 (132) | 35.09 (93) | 23.08 (3) | 42.86 (6) |
| Once | 9.67 (65) | 8.95 (34) | 10.94 (29) | 0.00 (0) | 14.29 (2) |
| Never | 31.55<br>(212) | 30.53 (116) | 36.23 (96) | 0.00 (0) | 0.00 (0) |
| Refused | 2.53 (17) | 2.37 (9) | 2.26 (6) | 15.38 (2) | 0.00 (0) |
| Saw/heard parent/household member in your home being hit/cut with an object (e.g., stick, |  |  |  |  |  |
| bottle, knife) |  |  |  |  |  |
| Many times | 19.64<br>(132) | 21.58 (82) | 13.96 (37) | 53.85 (7) | 42.86 (6) |
| A few times | 27.68<br>(186) | 26.84 (102) | 29.43 (78) | 23.08 (3) | 21.43 (3) |
| Once | 14.29 (96) | 13.95 (53) | 13.96 (37) | 7.69 (1) | 35.71 (5) |
| Never | 35.57<br>(239) | 35.26 (134) | 39.62 (105) | 0.00 (0) | 0.00 (0) |
| Refused |  | 2.37 (9) | 3.02 (8) | 15.38 (2) | 0.00 (0) |
| **One or no parents, parental separation, or divorce** |  |  |  |  |  |
| Parents ever separated or divorced |  |  |  |  |  |
| Yes | 50.45<br>(339) | 52.63 (200) | 45.66 (121) | 69.23 (9) | 64.29 (9) |
| No | 40.18<br>(270) | 38.16 (145) | 45.28 (120) | 23.08 (3) | 14.29 (2) |
| Not applicable | 9.38 (63) | 9.21 (35) | 9.06 (24) | 7.69 (1) | 21.43 (3) |
| Did your mother, father or guardian die? |  |  |  |  |  |
| Yes | 47.62<br>(320) | 47.89 (182) | 46.42 (123) | 53.85 (7) | 57.14 (8) |
| No | 50.74<br>(341) | (50.0 (190) | 52.45 (139) | 46.15 (6) | 42.86 (6) |
| Don't know/not sure/refused to answer | 1.64 (11) | 2.11 (8) | 1.13 (3) | 0.00 (0) | 0.00 (0) |
| **Parents** |  |  |  |  |  |
| **Emotional neglect** |  |  |  |  |  |
| Parents/guardians really understand your problems and worries |  |  |  |  |  |
| Always | 32.89<br>(221) | 34.47 (131) | 32.83 (87) | 0.00 (0) | 21.43 (3) |
| Most of the time | 16.96<br>(114) | 16.32 (62) | 18.49 (49) | 7.69 (1) | 14.29<br>(2) |
| Sometimes | 24.70<br>(166) | 22.89 (87) | 26.79 (71) | 23.08 (3) | 35.71<br>(5) |
| Rarely | 10.86 (73) | 8.68 (33) | 12.45 (33) | 38.46 (5) | 14.29<br>(2) |
| Never | 12.50 (84) | 15.26 (58) | 7.55 (20) | 30.77 (4) | 14.29<br>(2) |
| Refused | 2.08 (14) | 2.37 (9) | 1.89 (5) | 0.00 (0) | 0.00 (0) |
| Parents/guardians really knew what you were doing with your free time when not in school/work |  |  |  |  |  |
| Always | 26.34<br>(177) | 27.89 (106) | 25.66 (68) | 0.00 (0) | 21.43 (3) |
| Most of the time | 14.88<br>(100) | 16.58 (63) | 13.21 (35) | 0.00 (0) | 14.29 (0) |
| Sometimes | 22.92 | 21.84 (83) | 23.77 (63) | 30.77 (4) | 28.57 (4) |
|  | (154) |  |  |  |  |
| Rarely | 12.80 (86) | 10.00 (38) | 14.72 (39) | 46.15 (6) | 21.43 (3) |
| Never | 20.98 (141) | 21.32 (81) | 20.75 (55) | 23.08 (3) | 14.29 (2) |
| Refused | 2.08 (14) | 2.37 (9) | 1.89 (5) | 0.00 (0) | 0.00 (0) |
| **Physical neglect** |  |  |  |  |  |
| Parents/guardians not give you enough food even when they could easily have done so |  |  |  |  |  |
| Many times | 16.67 (112) | 18.16 (69) | 14.72 (39) | 7.69 (1) | 21.43 (3) |
| A few times | 33.18 (223) | 35.53 (135) | 29.81 (79) | 23.08 (3) | 42.86 (6) |
| Once | 13.10 (88) | 11.05 (42) | 14.34 (38) | 38.46 (5) | 21.43 (3) |
| Never | 35.12 (236) | 33.68 (128) | 38.49 (102) | 30.77 (4) | 14.29 (2) |
| Refused | 1.93 (13) | 1.58 (6) | 2.64 (7) | 0.00 (0) | 0.00 (0) |
| Parents/guardians were too drunk or intoxicated by drugs to take care of you |  |  |  |  |  |
| Many times | 11.61 (78) | 14.74 (56) | 6.42 (17) | 0.00 (0) | 35.71 (5) |
| A few times | 16.82 (113) | 17.11 (65) | 15.09 (40) | 23.08 (3) | 35.71 (5) |
| Once | 6.99 (47) | 6.58 (25) | 6.04 (16) | 38.46 (5) | 7.14 (1) |
| Never | 61.61 (414) | 58.16 (221) | 69.81 (185) | 38.46 (5) | 21.43 (3) |
| Refused | 2.98 (20) | 3.42 (13) | 2.64 (7) | 0.00 (0) | 0.00 (0) |
| Parents/guardians not send you to school even when it was available |  |  |  |  |  |
| Many times | 20.09 (135) | 23.68 (90) | 14.72 (39) | 15.38 (2) | 28.57 (4) |
| A few times | 24.11 (162) | 26.32 (100) | 17.74 (47) | 46.15 (6) | 64.29 (9) |
| Once | 12.80 (86) | 11.84 (45) | 15.09 (40) | 7.69 (1) | 0.00 (0) |
| Never | 40.33 (271) | 35.00 (133) | 50.19 (133) | 30.77 (4) | 7.14 (1) |
| Refused | 2.68 (18) | 3.16 (12) | 2.26 (6) | 0.00 (0) | 0.00 (0) |
| **Violence outside the household** |  |  |  |  |  |
| **Bullying** |  |  |  |  |  |
| How often were you bullied |  |  |  |  |  |
| Many times | 51.49 (346) | 46.58 (177) | 54.34 (144) | 100.0 (13) | 85.71 (12) |
| A few times | 24.70 (166) | 27.63 (105) | 22.64 (60) | 0.00 (0) | 7.14 (1) |
| Once | 4.32 (29) | 3.95 (15) | 5.28 (14) | 0.00 (0) | 0.00 (0) |
| Never | 14.73 (99) | 16.58 (63) | 13.21 (35) | 0.00 (0) | 7.14 (1) |
| Refused | 4.76 (32) | 5.26 (20) | 4.53 (12) | 0.00 (0) | 0.00 (0) |
| **Community violence** |  |  |  |  |  |
| Saw or heard someone being beaten up in real life |  |  |  |  |  |
| Many times | 65.03<br>(437) | 63.95 (243) | 65.28 (173) | 76.92 (10) | 78.57<br>(11) |
| A few times | 25.15<br>(169) | 25.00 (95) | 25.66 (68) | 23.08 (3) | 21.43 (3) |
| Once | 2.98 (20) | 2.63 (10) | 3.77 (10) | 0.00 (0) | 0.00 (0) |
| Never | 5.06 (34) | 5.79 (22) | 4.53 (12) | 0.00 (0) | 0.00 (0) |
| Refused | 1.79 (12) | 2.63 (10) | 0.75 (2) | 0.00 (0) | 0.00 (0) |
| Saw or heard someone being stabbed or shot in real life |  |  |  |  |  |
| Many times | 37.65<br>(253) | 37.37 (142) | 34.34 (91) | 69.23 (9) | 78.57<br>(11) |
| A few times | 24.85<br>(167) | 23.95 (91) | 27.92 (74) | 7.69 (1) | 7.14 (1) |
| Once | 9.38 (63) | 7.89 (30) | 11.32 (30) | 15.38 (2) | 7.14 (1) |
| Never | 25.15<br>(169) | 27.11 (103) | 24.15 (64) | 7.69 (1) | 7.14 (1) |
| Refused | 2.98 (20) | 3.68 (14) | 2.26 (6) | 0.00 (0) | 0.00 (0) |
| See or hear someone being threatened with a knife or gun in real life |  |  |  |  |  |
| Many times | 52.08<br>(350) | 50.26 (191) | 52.08 (138) | 76.92 (10) | 78.57 (11) |
| A few times | 25.74<br>(173) | 25.79 (98) | 26.79 (71) | 15.38 (2) | 14.29 (2) |
| Once | 6.55 (44) | 6.58 (25) | 6.79 (18) | 7.69 (1) | 0.00 (0) |
| Never | 13.84 (93) | 15.53 (59) | 12.45 (33) | 0.00 (0) | 7.14 (1) |
| Refused | 1.79 (12) | 1.84 (7) | 1.89 (5) | 0.00 (0) | 0.00 (0) |
| **Collective Violence** |  |  |  |  |  |
| Forced to go and live in another place |  |  |  |  |  |
| Many times | 34.52<br>(232) | 38.68 (147) | 29.06 (77) | 23.08 (3) | 35.71 (5) |
| A few times | 21.43<br>(144) | 17.11 (65) | 26.04 (69) | 53.85 (7) | 21.43 (3) |
| Once | 17.71<br>(119) | 17.11 (65) | 18.87 (50) | 7.69 (1) | 21.43 (3) |
| Never | 24.85<br>(167) | 25.53 (97) | 24.53 (65) | 15.38 (2) | 21.43 (3) |
| Refused | 1.49 (10) | 1.58 (6) | 1.51 (4) | 0.00 (0) | 0.00 (0) |
| Experience the deliberate destruction of your home |  |  |  |  |  |
| Many times | 22.92<br>(100) | 27.11 (103) | 16.60 (44) | 23.08 (3) | 28.57 (4) |
| A few times | 8.33 (56) | 8.16 (31) | 7.17 (19) | 23.08 (3) | 21.43 (3) |
| Once | 17.11 | 16.58 (63) | 18.87 (50) | 15.38 (2) | 0.00 (0) |
|  | (115) |  |  |  |  |
| Never | 49.26 (331) | 45.79 (174) | 54.72 (145) | 38.46 (5) | 50.00 (7) |
| Refused | 2.38 (16) | 2.37 (9) | 2.64 (7) | 0.00 (0) | 0.00 (0) |
| Beaten up by soldiers, police, militia, or gangs |  |  |  |  |  |
| Many times | 11.46 (77) | 11.84 (45) | 8.68 (23) | 38.46 (5) | 28.57 (4) |
| A few times | 5.51 (37) | 5.53 (21) | 4.91 (13) | 23.08 (3) | 0.00 (0) |
| Once | 20.68 (139) | 18.42 (70) | 23.77 (63) | 23.08 (3) | 21.43 (3) |
| Never | 60.71 (408) | 62.63 (238) | 61.13 (162) | 7.69 (1) | 50.00 (7) |
| Refused | 1.64 (11) | 1.58 (6) | 1.51 (4) | 7.69 (1) | 0.00 (0) |
| Family member or friend killed or beaten up by soldiers, police, militia, or gangs |  |  |  |  |  |
| Many times | 13.24 (89) | 12.89 (49) | 12.45 (33) | 30.77 (4) | 21.43 (3) |
| A few times | 10.86 (73) | 10.00 (38) | 12.45 (33) | 15.38 (2) | 0.00 (0) |
| Once | 24.55 (165) | 21.05 (80) | 27.92 (74) | 38.46 (5) | 42.86 (6) |
| Never | 48.81 (328) | 53.42 (203) | 44.91 (119) | 7.69 (1) | 35.71 (5) |
| Refused | 2.53 (17) | 2.63 (10) | 2.26 (6) | 7.69 (1) | 0.00 (0) |

Note. \*One person did not respond to this question and was dropped.

### Abuse (Physical, Emotional, Sexual)

According to respondents’ self-report, prevalence of physical, emotional, and sexual abuse was high in the full sample (64% to 74%, 63% to 73%, and 20% to 43%, respectively). However, notable differences in frequency and severity emerged by self-identification categories. A higher percentage of self-identified homosexuals and lesbians endorsed ‘many times’ for all types of abuse compared to self-identifying women or men. For instance, about two-third of self-identified homosexuals (61.54%) and one-third of self-identified lesbians (35.17%) said a parent/household member had hit/cut them many times, compared to 26.84% of self-identified women and 21.89% of self-identified men. A substantial majority of self-identified lesbians (85.71%) and more than two-third of self-identified homosexuals (69.23%) reported threat or actual abandonment by a parent/household member many times, compared to less than one-third of self-identified women (29.47%) and one-fifth of self-identified men (18.49%). All sexual abuse items elicited similar response patterns including forced intercourse. More than two-third (69.23%) of self-identified homosexuals and about three-fifth of self-identified lesbians (57.14%) had been subjected to oral/anal/vaginal intercourse many times, compared to 5% and 1.13% of self-identified women and men respectively.

### Family Dysfunction

Prevalence of family dysfunction was highest for individuals self-identifying as homosexual or lesbian. In comparison to self-identified women or men, much higher percentages of self-identified homosexuals and lesbians reported living with an alcoholic/drug abusing household member (49.74% and 43.02% versus 92.31% and 85.71% respectively), and with a household member who was mentally ill (35.53% and 35.47% versus 53.85% and 71.43% respectively). Over half of self-identified homosexuals (53.85%) and about two-fifth of self-identified lesbians (42.86%) had witnessed a family member get hit or cut many times (versus 21.58% self-identified women and 13.96% self-identified men).

### Neglect by Parents

The findings related to neglect as an ACE were mixed. For instance, about two-thirds of self-identified homosexuals (61.54%) and three-fourth of self-identified lesbians (78.57%) revealed that at least one time a parent/guardian was too intoxicated to be able to take care of them (versus 38.42% self-identified women, 27.55% self-identified men). In relation to food deprivation even when food is available, 85.7% self-identified lesbians had experienced that at least once. Such neglect was also reported by 69% self-identified homosexuals, closely followed by self-identified women at 64.42% and self-identified men at 58.36%. As high as 93% of self-identified lesbians (93%), two-third of self-identified homosexuals (61.43%) and about half of self-identified women (46.84%) said at least one time a parent/guardian had not sent them to school even when available compared to one-third of self-identified men (30.20%).

### Violence Outside the Household

Prevalence of bullying was widespread (81% of the full sample reported at least one victimization). Notably, all self-identified homosexuals and 86% of self-identified lesbians had experienced frequent bullying.

Consistent with extant knowledge of interpersonal violence in Cité Soleil, most respondents reported often witnessing violence such as seeing or hearing community members getting beaten up (65.03%), threatened with a knife/gun (52.08%), or stabbed/shot (37.65%). However, percentages were highest for self-identified homosexuals who reported often seeing or hearing community members getting beaten up (76.92%), threatened with a knife/gun (76.92%), or stabbed/shot (69.23%) as well as lesbians (78.57% often for all three events).

A substantial percentage of our full sample reported forced relocation (73.66%). However, consistent with previous findings (Joshi et al., 2021), women relocated more frequently, followed by self-identified lesbians, men, and homosexuals (38.68%, 35.71%, 29.06%, and 23.08% respectively). Further, about 38% of the full sample said they endured beatings from soldiers, police, militia, and/or gangs. Again, a higher percentage of self-identified homosexuals and lesbians reported such beatings ‘many times’ (38.46%, 28.57% respectively) in comparison to 11.84% self-identified women and 8.68% self-identified men.

## Discussion

We set out to assess ACEs in Haiti’s Cité Soleil and to intentionally include underrepresented SGM. We found a high prevalence of physical, emotional, and sexual abuse in the full sample. However, a higher percentage of SGM subsample reported a higher frequency and severity of ACEs than others. On all items related to interpersonal violence during childhood, participants in the SGM sub sample reported disproportionate experiences. In addition, SGM participants reported not finding understanding or concern from parents, being deprived of basic needs, forced relocation, bullying and other forms of violence outside the home. In addition, the finding, despite the small SGM sub-sample, that a higher percentage of self-identified homosexuals and lesbians endorsed ‘many times’ for all items of abuse compared to others suggests a need for further research using a contextually grounded but evidence-informed version of the ACE-IQ. This is salient in view of extant knowledge regarding adverse biopsychosocial impacts of ACEs on individuals and on the additional distress experienced by Haitian SGM. Our findings have significant implications for trauma symptoms, depression, anxiety, and other psychological distress, especially given the known health and mental health disparities between Haitians and others in the Caribbean region, and health disparities between SGM and others globally (Lund & Burgess, 2021).

### Limitations

This study used non-probability sampling methods in a cross-sectional survey; hence, findings cannot be generalized to Haiti or beyond the study timeframe. Our participants self-reported on situations that happened before the age of 18; hence, data were subject to recall bias. Data collectors reported accommodating participants who could not read by administering the survey to them, which is how the ACE-IQ should have been administered to all participants. Despite its limitations, this study is an important first step in documenting potentially disproportionate burden of ACEs experienced by SGM in Haiti’s Cite Soleil, and a compelling reason to further sexual-diverse and gender-diverse research in LIC/LMICs such as Haiti.

## Conclusion

Our findings reconfirm Sharma’s 1986 assertation that measures of social or psychological functioning are:

> …affected in a variety of ways by political, social, and philosophical considerations [and] personal attitudes, often emotionally laden, tend to color measurement efforts…[Thus]…representatives of the minority groups, with an understanding of the testing and measurement requirements of their groups, should be involved in the [development] (p. 122)

Such involvement might enable more distinct and comprehensive categories to disentangle sex, gender, and sexual orientation as well as self-identified roles and preferences, which would be beneficial in future iterations of the ACE-IQ. For example, in this study, in the absence of an ‘Other’ category, the interviewer perhaps would have recorded 19 individuals as either male, female or would have left it blank. Further, for the 10 participants with no record, perhaps, the interviewer was either unsure or indeed missed the item. Further, the self-identification item did not allow for ‘Check all that apply’; thus, gender and sexual orientation categories were conflated, and they were forced to either pick a gender or a sexual orientation, not both. Also noteworthy is that no participants reported ‘I have a different identification.’ Perhaps, the participants were either satisfied with response categories offered or were unaware of alternative terminologies that can be used to identify one’s gender and sexual orientation.

The question on sex and gender in an instrument such as the ACE-IQ, which is often used in LMICs, may need to include a brief paragraph which details distinctions among sex, gender, sexual orientation, and cultural roles assumed by SGM in various contexts. This would provide a unique opportunity to: (1) Increase knowledge among Haiti’s SGM about available options for self-classification, and (2) Reject contextual norms that identify them in terms of sexual orientation rather than gender identity. For example, the ACE-IQ item ‘Sex as observed/recorded’ was difficult in this context and data collectors reported leaving that item blank since the meaning ‘of observed’ was not clear. They indicated that in Haiti, individuals are categorized based on social roles, which in turn is based on individuals’ manner of dress, gender expression or perceived sexual orientation. Finally, our colleagues clarified that in Haiti, the terms male and female describe animals for reproductive purposes and that the terms masculine or feminine are used to describe men and women. Thus, context and language may need to be considered when administering the ACE-IQ where individuals’ roles take precedence over their sex or self-identification (Rahill et al., 2021).

To our knowledge, this is among the first ACE-IQ administrations in Haiti, and the first to intentionally include SGM and to consider self-identification in a community survey. This study offers several lessons for the field in terms of conducting equitable research that underscores the importance of inclusiveness of different populations (e.g., how to frame questions that offer SGM more freedom to answer questions related to gender, gender identity, gender expression, sexual orientation; role of context in collecting data using ACE-IQ). Notably, although ACEs can have devastating outcomes when positive childhood experiences (PCEs) are few or absent, PCEs can serve as protective factors against adverse adult health outcomes (Children’s Bureau, 2020). Interventions that provide PCEs for Cité Soleil residents, with particular emphasis on those whose SGM identities are emerging, might mitigate adverse biopsychosocial experiences across the lifespan.

## Data Availability

All data produced in the present study are available upon reasonable request to the authors

